# Long-term air pollution and other risk factors associated with COVID-19 at the census-tract-level in Colorado

**DOI:** 10.1101/2021.02.19.21252019

**Authors:** Kevin Berg, Paul Romer Present, Kristy Richardson

## Abstract

An effective response to the COVID-19 pandemic requires identification of the factors that affect the severity and mortality of the disease. Previous nationwide studies have reported links between long-term PM2.5 concentrations and COVID-19 infection and mortality rates. In order to translate these results to the state level, we use Bayesian hierarchical models to explore potential links between long-term PM2.5 concentrations and census tract-level rates of COVID-19 outcomes (infections, hospitalizations, and deaths) in Colorado. We explicitly consider how the uncertainty in PM2.5 estimates affect our results by comparing four different PM2.5 surfaces from academic and governmental organizations. After controlling for 20 census tract level covariates including race/ethnicity, socioeconomic status, social distancing, age demographics, comorbidity rates, meteorology, and testing rate, we find that our results depend heavily on the choice of PM2.5 surface. Using PM2.5 estimates from the United States EPA, we find that a 1 µg/m^3^ increase in long term PM2.5 is associated with a statistically significant 25% increase in the relative risk of hospitalizations and a 35% increase in mortality. Results for all other surfaces and outcomes were not statistically significant. At the same time, we find a clear association between communities of color and COVID-19 outcomes at the Colorado census-tract level that is minimally affected by the choice of PM2.5 surface. A per-interquartile range (IQR) increase in the percent of non-African American people of color was associated with a 31%, 44%, and 59% increase in the relative risk of infection, hospitalization, and mortality respectively, while a per-IQR increase in the proportion of non-Hispanic African Americans was associated with a 4% and 7% increase in the relative risk of infections and hospitalizations. These results have strong implications for the implementation of an equitable public health response during the crisis and suggest targeted areas for additional air monitoring in Colorado.

## Introduction

Although a full accounting of the biological and societal risk factors driving the spread of the SARS-CoV-2 virus will require years of study to be fully understood, several recent epidemiological studies have provided national assessments of the role of important risk factors of COVID-19-related morbidity and mortality. In particular, a growing body of evidence points to fine particulate matter (PM2.5) air pollution exposure and racial/ethnic disparities as being possible drivers of increased vulnerability to complications related to COVID-19. The Harvard School of Public Health released a well-publicized study in April that found an increase of 1 microgram per cubic meter (µg/m^3^) in long-term average PM2.5 concentration was associated with an 11% increase in the COVID-19 mortality rate at the U.S. county level (Wu et al. 2020). However, estimated effects for the same increase in PM2.5 reported elsewhere have ranged from a 4% decrease (Travaglio et al., 2021) to a 17% increase (Tian et al., 2020). Further, there is no one universally accepted standard for measuring community-scale exposure to PM2.5 air pollution, and studies to date have not compared effect estimates reported from different PM2.5 exposure surfaces. In addition, to overcome incomplete reporting of COVID-19 morbidity and mortality by race/ethnicity, two recent national studies analyzed publicly available data from U.S. counties to describe racial/ethnic disparities for non-Hispanic African American and Hispanic/Latino populations, respectively (Millet et al., 2020; Rodriguez-Diaz et al., 2020). Nationwide, these studies reported that counties with a greater percentage of non-Hispanic African American and Hispanic/Latino residents have statistically higher COVID-19 burdens. However, while these recent U.S.-based studies of COVID-19 risk factors accounted for a variety of confounding variables, they all suffered from relatively coarse spatial resolution by relying on county-level data. This limits the ability of state and local public health officials to translate these findings for decision making within their own jurisdictions.

This analysis builds on these previous studies by constructing a statewide census tract-level analysis in the U.S. state of Colorado. We fit spatial regression models of census tract-level counts of three COVID-19 outcomes (infection, hospitalization, and mortality) using Bayesian hierarchical random effects models. For the main analysis, we considered four different measures of PM2.5 air pollution exposure, along with additional census tract-level confounding factors including race/ethnicity, socioeconomic status, multiple measures of social distancing, age demographics, rates of comorbidity prevalence and hospitalization, time elapsed since the first reported case, nearby hospital capacity, and overall testing rate. The full model uses random effects to account for spatially correlated as well as uncorrelated variation across all census tracts, and includes a county-level random effect to account for uncorrelated variation across counties.

## Methods

### COVID-19 Outcome Data

We obtained census tract-level counts of COVID-19 cases, hospitalizations, and deaths from the Colorado Electronic Disease Reporting System (CEDRS) as well as tract-level counts of unique individuals tested for COVID-19 from Colorado’s electronic lab reporting (ELR) system. All counts were stratified by age group and gender, and prior to aggregation to census tract, individual records were geocoded using the address information provided within the CEDRS and ELR databases. While CDPHE ascertains patient address information for COVID-19 cases within the CEDRS database, missing address information for individuals receiving a COVID-19 test within the ELR database can be filled in either by matching to other surveillance data sources or substituted by the address of the health care provider. In all, approximately 88% of cases and 93% individuals with a COVID-19 test were geocoded to a census tract. For our main analysis, we computed the total number of all COVID-19 infections, hospitalizations, and deaths by census tract between March 1, 2020 and August 31, 2020. The COVID-19 data were pulled from the CEDRS and ELR databases on October 22, 2020. Figure 1 displays the crude rate of each outcome per 100,000 population by county in Colorado.

**Figure 1:**
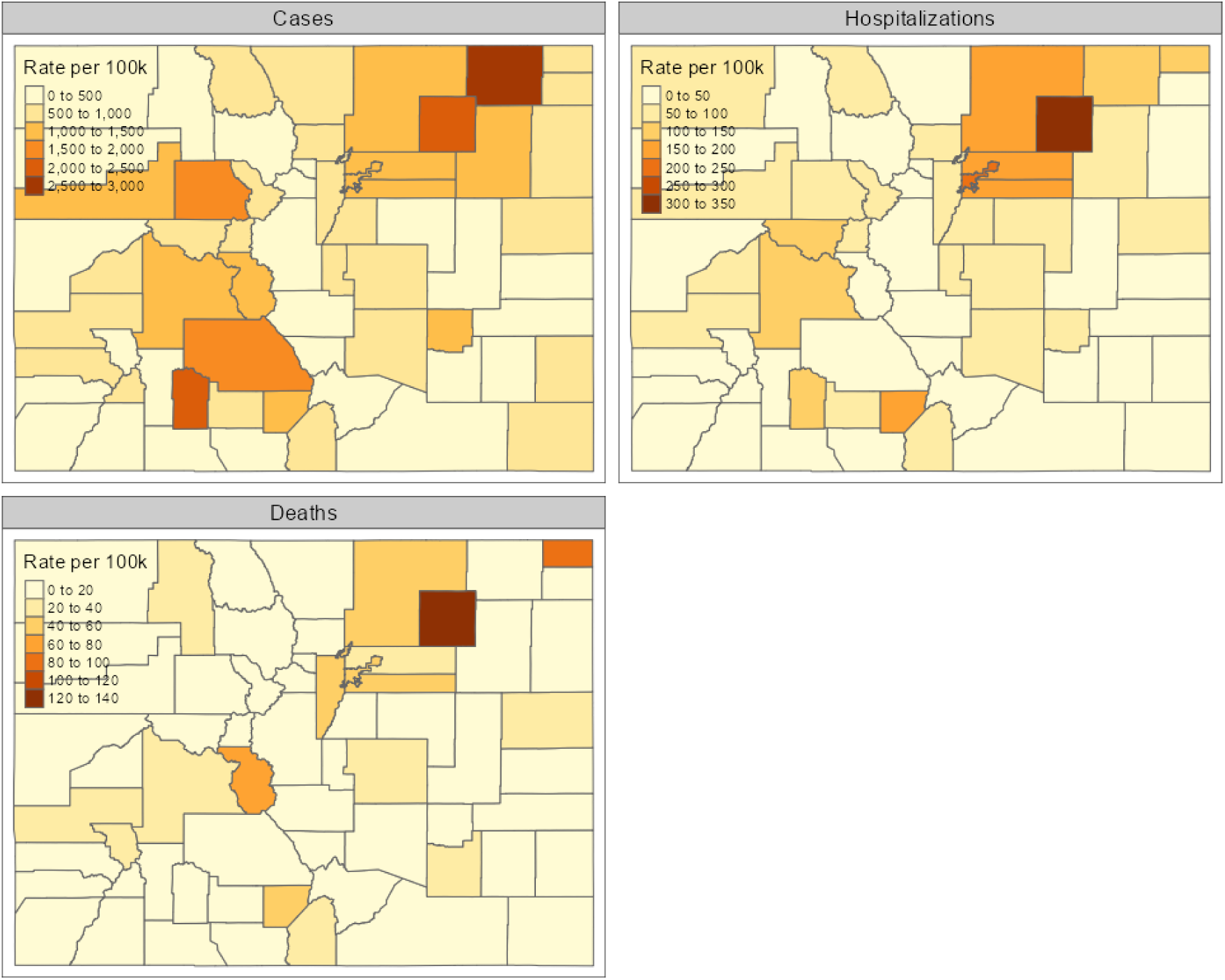
COVID-19 outcomes by county, 1 Mar - 31 August 2020.

### PM2.5 Pollution Data

The most widely accepted measurements of PM2.5 come from regulatory-grade measurements reported to the US Environmental Protection Agency (EPA). However, while these monitors are excellent at capturing regional air quality, these networks are too coarse to accurately characterize PM2.5 gradients within cities, which can vary sharply on spatial scales of 1 km or less (Karner et al., 2010; Robinson et al., 2018). In addition, many rural areas, including a significant portion of Colorado, are 50 km or more from the nearest monitor (Technical Services Program, 2020) To overcome these limitations, several research groups and government agencies have developed estimated PM2.5 surfaces that combine EPA monitoring data with other sources of information, such as satellite measurements of aerosol optical depth, land-use information, or the output from chemical transport models (Diao et al. 2019). We identified four different PM2.5 surfaces for use in our analysis. Details about the different surfaces are presented in Table 1.

**Table 1:** Characteristics of the PM2.5 surfaces used in this analysis

| Abbreviated Name | Creator | Time Period | Spatial Resolution | Data Sources Included | Reference |
| --- | --- | --- | --- | --- | --- |
| ACAG | Atmospheric Composition Analysis Group at Washington University | 2000 - 2018 | 0.01° x 0.01° (Approximately 1 km x 1 km) | EPA monitors, computer modeling (GEOS-Chem), satellite measurements | van Donkelaar et al., 2019 |
| EPA | Environmental Protection Agency | 2002 - 2017 | Census tract | EPA monitors, computer modeling (CMAQ) | Berrocal et al., 2012 |
| Pierce | Pierce Group at Colorado State University | 2006 - 2018 | 15 km x 15 km | EPA monitors | O'Dell et al., 2019 |
| Schwartz | Schwartz Group at Harvard University | 2000 - 2016 | 1 km x 1 km | EPA monitors, computer modeling (CMAQ), satellite measurements, land-use | Di et al., 2019 |

All four surfaces considered in this analysis are available over the entire continental United States. Three of them (ACAG, EPA, and Pierce) are publicly available. The fourth (Schwartz) was provided upon request. The EPA surface was provided at the census-tract level, and no additional processing was necessary. We transformed the gridded products into census tract-level averages by assuming the PM2.5 concentration was constant across each grid cell and using an area-weighted average. Figure 2 shows the long-term average PM2.5 concentrations for each surface over the entire state of Colorado (panels a-d) and over the Denver metro area (panels e-h). While all four surfaces identify Denver and the Front Range region as the areas with the highest average concentrations of PM2.5, there are significant differences among the four surfaces, particularly over the Rocky Mountains and in the area to the northeast of Denver.

**Figure 2:**
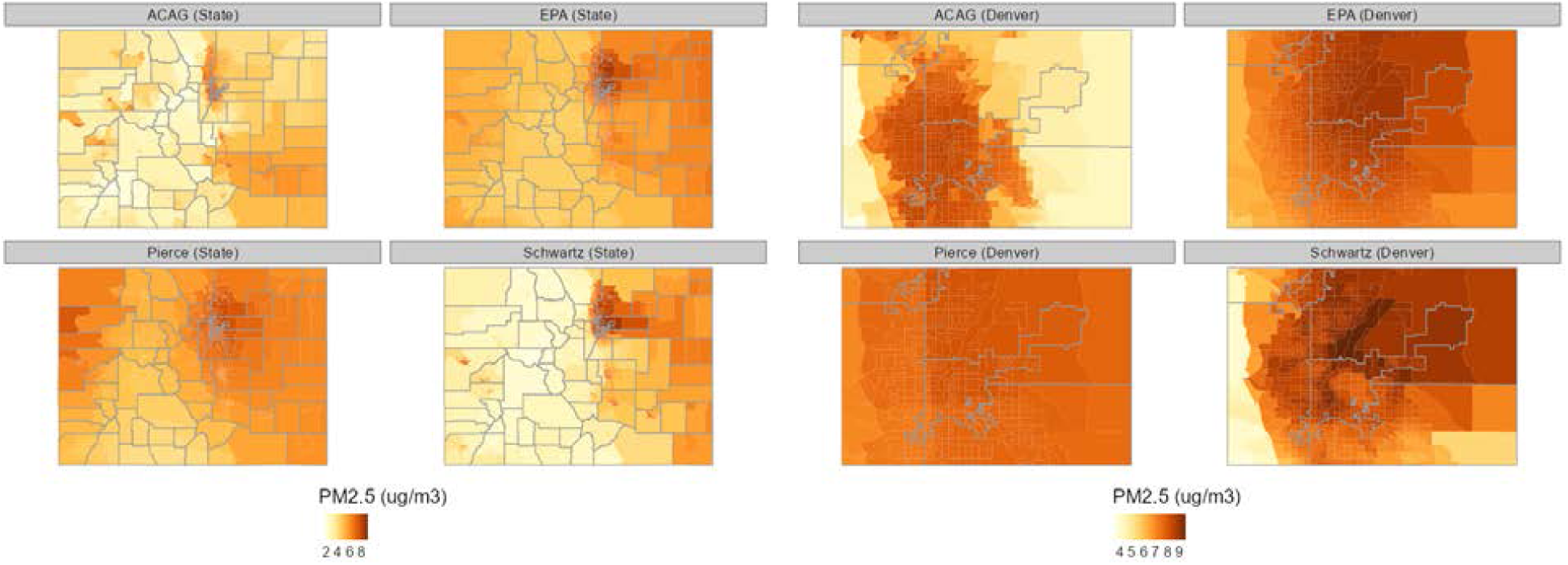
Comparison of the four different PM2.5 surfaces over the state of Colorado and in the Denver metro area.

### Potential Confounding Variables

We identified a total of 42 additional census tract-level factors based on those included as confounders in two previous U.S. county-level analyses of the association between PM2.5 exposure and COVID-19 mortality (Wu et al. 2020; Knittel & Ozaltun, 2020). However, we detected significant problems with multicollinearity among several of these proposed covariates when we translated them to the census-tract level in Colorado. Therefore, we selected a reduced set of 20 covariates through stepwise regression and manual examination of the variance inflation factors (VIFs). Descriptions of all 42 potential confounding variables and details of the variable selection procedure are presented in the supplemental information.

The selected set of covariates includes six measures from the U.S. Census American Community Survey (ACS) 2014-2018 five-year estimates describing the age, race/ethnicity, and income distribution of each census tract. The race/ethnicity of each census tract was characterized by the proportion of the population that is non-Hispanic African American and the proportion that is non-African American people of color. The latter demographic group is composed mainly of Colorado’s Hispanic/Latino community, but also includes the proportion that is American Indian and Alaska Native, Asian, Native Hawaiian and other Pacific islander, and two or more races. We included three measures describing the age distribution in each census tract: the proportion of the population between 18 and 44 years old, the proportion between 45 and 64 years old, and the proportion over 64 years old. We also included the percent of census tract population living in households with family income in the last 12 months below the federal poverty line.

Because COVID-19 transmission is thought to be affected by meteorology (Merow and Urban, 2020; Poirier et al., 2020), we included the 30-year average summer temperature and summer relative humidity at the census-tract level as covariates.

We include five measures from the ACS related to social distancing: population density, the proportion of the population living in overcrowded housing, the proportion employed in an essential industry, and the proportion living in correctional facilities, nursing homes, or mental hospitals. We also considered two measures of social distancing derived from mobile phone data provided by the SafeGraph COVID-19 Data Consortium and compiled for the Colorado Department of Public Health and Environment (CDPHE) by Citizen Software Engineers. The first is a derived measure of virus exposure combining average daily travel to a census tract with the infection rate of the corresponding county of origin. Higher measures correspond to greater volumes of travel to a census tract originating in counties with higher COVID-19 infection rates. The second is an index of the average daily amount of time spent at home per census tract derived from mobile phone location data. For the main analysis, we averaged these variables over the time period spanning March 1, 2020 to the end of the City and County of Denver’s stay-at-home order on May 8, 2020.

We also included two measures accounting for underlying health factors in each census tract. These measures were based on estimates of census tract level adult prevalence of diabetes, heart disease, obesity and smoking and the age-adjusted rates of hospitalization for asthma, diabetes, heart disease, and influenza. The prevalence estimates were derived from multiple years of Colorado Behavioral Risk Factor Surveillance System data (2014-2017), and the hospitalization data was computed from 5 years (2014-2018) of discharge data from the Colorado Hospital Association. We used principal component analysis to reduce the comorbidity measures to a pair of uncorrelated principal components. The first comorbidity component was strongly correlated with higher prevalence rates while the second comorbidity component was strongly correlated with higher age-adjusted hospitalization rates.

Next, to control for differential testing patterns, we considered the crude testing rate per census tract population from the 2014-2018 ACS estimates. We also considered the time elapsed in days since the first case was reported for each census tract, as well as the number of certified hospital beds per unit population to control for differences due to timing of initial outbreak and overall health care access.

### Statistical Analysis

We applied a Bayesian hierarchical random effects model, based on a design first proposed by Besag, York and Mollie (BYM) (Besag et al., 1991), to investigate the association between long-term PM2.5 concentrations and counts of COVID-19 outcomes at the census-tract level. The BYM model includes both an intrinsic conditional auto-regressive (ICAR) component to account for spatial autocorrelation between census tracts and an exchangeable random-effect component to account for uncorrelated variation across all census tracts. The prior distributions controlling for the variance of the ICAR and exchangeable components were selected to give approximately equal emphasis to the two components (Bernardinelli et al., 1995; Morris et al., 2019). Our model also includes a county-level exchangeable random-effect component to account for uncorrelated variation due to unmeasured county-level factors. Our primary analyses included 12 separate models, one for each combination of three COVID-19 outcomes and four PM2.5 exposure surfaces. In each model, the census tract-level count of each COVID-19 outcome was assumed to follow a Poisson distribution using the indirectly age-sex standardized expected count as an offset term.

We adjusted each of our 12 models by the same set of 20 census tract-level covariates described in the previous section. PM2.5 measurements in this analysis represent long-term annual average census-tract concentrations reported in units of µg/m^3^. Hence, our reported regression coefficients for the PM2.5 variables represent the effect of a 1 µg/m^3^ increase in long-term annual average PM2.5 concentration at the census-tract level. Further, we scaled the two race/ethnicity variables by their respective interquartile ranges (IQR), and thus reported regression coefficients compare a census-tract in the 75th-percentile of a given race/ethnicity variable to a census-tract in the 25th-percentile. In other words, it compares a census-tract with a typical high proportion of either non-Hispanic African Americans or non-African American people of color to a census-tract with a typical low proportion. All other variables were converted to z-scores and their coefficients represent the effect of a one-standard-deviation increase above their respective means.

### Sensitivity Analysis

For each of our 12 models, we conducted four main sensitivity analyses. First, to assess the impact of our choice of offset term, we explored an alternative offset term that used the census-tract population per 100,000 from the American Community Survey 2014-2018 5-year estimates. Second, while including spatially correlated random effects often leads to more accurate estimates of fixed effects (Beale et al., 2010), it has also been suggested that their inclusion may introduce bias in the estimates of the fixed effects (Hodges & Reich, 2010). To account for the latter possibility, we created a separate set of models by replacing the BYM component with a census tract-level random intercept term. Third, to assess robustness of the results to study area selection, we stratified census tracts into two groups, the first including only census tracts that fall within the 5 most populous counties in the Denver metro area (Adams, Arapahoe, Denver, Douglas, and Jefferson Counties), and the second including all tracts outside of these counties. The purpose was to rule out the possibility that any apparent associations observed in the main analyses were due only to the strong influence of high-population density census tracts in the state’s major population centers. Finally, to assess the sensitivity of observed associations during different phases of the pandemic, we disaggregated the analysis into 3 time periods: a period spanning March 1, 2020 to the end of Denver’s stay at home order on May 8, 2020, a subsequent period spanning May 9, 2020 to June 30, 2020, and a third period spanning July 1, 2020 to August 31, 2020. Results of our study area and time period sensitivity analysis are provided in our supplementary materials.

### Model Selection

Considering our main analysis together with our sensitivity analyses of the choice of offset term and method of addressing spatial autocorrelation, our next task was to select the best-fitting model among four candidate study designs. For convenience, we refer to the competing study designs as relative risk - BYM, relative risk - i.i.d., relative rate - BYM, and relative rate - i.i.d. Here, relative risk describes the use of the expected count as the offset term in the regression model whereas relative rate refers to the use of census-tract population per 100,000 in the offset term. The naming convention reflects the interpretation of the exponentiated regression coefficients, which in the former case can be interpreted as the multiplicative effect on the census-tract relative risk, while in the latter case would be interpreted as the multiplicative effect on the relative rate per 100,000 population. The i.i.d. model uses a tract-level random intercept term to account for uncorrelated variation across all census tracts, while the BYM models include both a random intercept and an ICAR component at the census-tract level. For each study design, we built a series of nested models, starting with a null model that included only the respective offset term and random effects as regressors. We then estimated separate unadjusted risk/rate ratios for the effect of our four different measures of long-term PM2.5 exposure on each COVID-19 outcome. We then estimated fully adjusted models including all other census-tract level characteristics to control for potential confounding of the PM2.5 effect.

We considered two primary factors to select an optimal model: the deviance information criterion (DIC) and the Moran’s I statistic of the model residuals. Briefly, the DIC is a measure of model fit that accounts for both goodness of fit and the effective number of parameters. Lower DIC values indicate better model fit, and differences in DIC greater than 10 are generally considered meaningful (Spiegelhalter et al. 2002). Moran’s I is a correlation coefficient that measures the overall spatial autocorrelation of the model residuals. The null hypothesis of the Moran’s I test states that the residuals are not spatially correlated, and thus a small p-value yields evidence that significant residual spatial correlation is present.

All analyses were conducted using the R statistical software environment version 3.6.2, and the main analysis was implemented in the R-INLA package version 19.09.03 (R Core Team, 2019; Rue et al., 2009). To protect privacy, COVID-19 outcomes at the census-tract level cannot be released publically; however, all code to complete this analysis can be made available upon request.

## Results

In this study, we analyzed counts of COVID-19 outcomes from 1237 populated census tracts in the U.S. state of Colorado. Between March 1 and August 31, 2020, the average Colorado census tract had accumulated 42.8 infections (range: 0 to 338), 5.1 hospitalizations (range: 0 to 58), and 1.4 deaths (range: 0 to 40), while 7 (0.56%), 175 (14.15%), and 637 (51.5%) tracts had counts of zero for each outcome, respectively. Meanwhile, pooling the four PM2.5 surfaces together, the average census tract had a long-term annual average PM2.5 exposure of 6.89 µg/m^3^ with a range between 1.78 and 9.95 µg/m^3^, depending on the surface source. Unadjusted risk/rate ratios from each model show a positive association between PM2.5 exposure and COVID-19 infection, hospitalization, and mortality, regardless of the provenance of the exposure surface (Figure 3).

**Figure 3:**
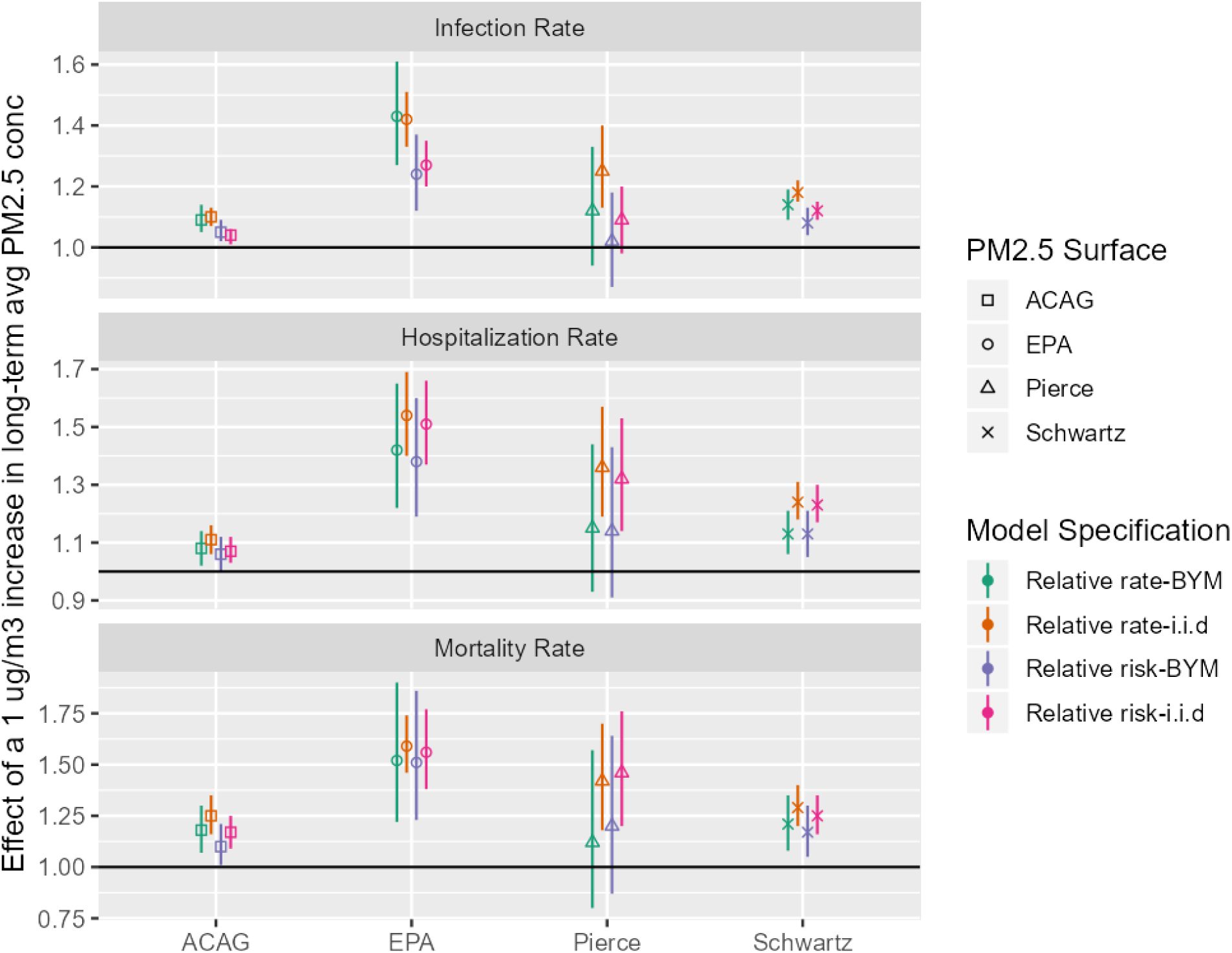
Unadjusted effect of a 1µg/m^3^ increase in long-term average PM2.5 concentration on three COVID outcomes, depending on the source of PM2.5 surface and model specification.

After adjusting for potential confounding variables, the analyses using the EPA, Pierce, or Schwartz surfaces typically show positive associations between PM2.5 exposure and COVID-19 infection, hospitalization, and mortality, while the analysis using the ACAG surface shows a negative association between PM2.5 exposure and all 3 COVID-19 outcomes (Figure 4). In both the adjusted and unadjusted models, the associations between long-term PM2.5 exposure and the three COVID-19 outcomes were relatively robust to the choice of study design. That is, for any given PM2.5 surface, the adjusted posterior mean for each measure of association was generally stable across each of the four study designs.

**Figure 4:**
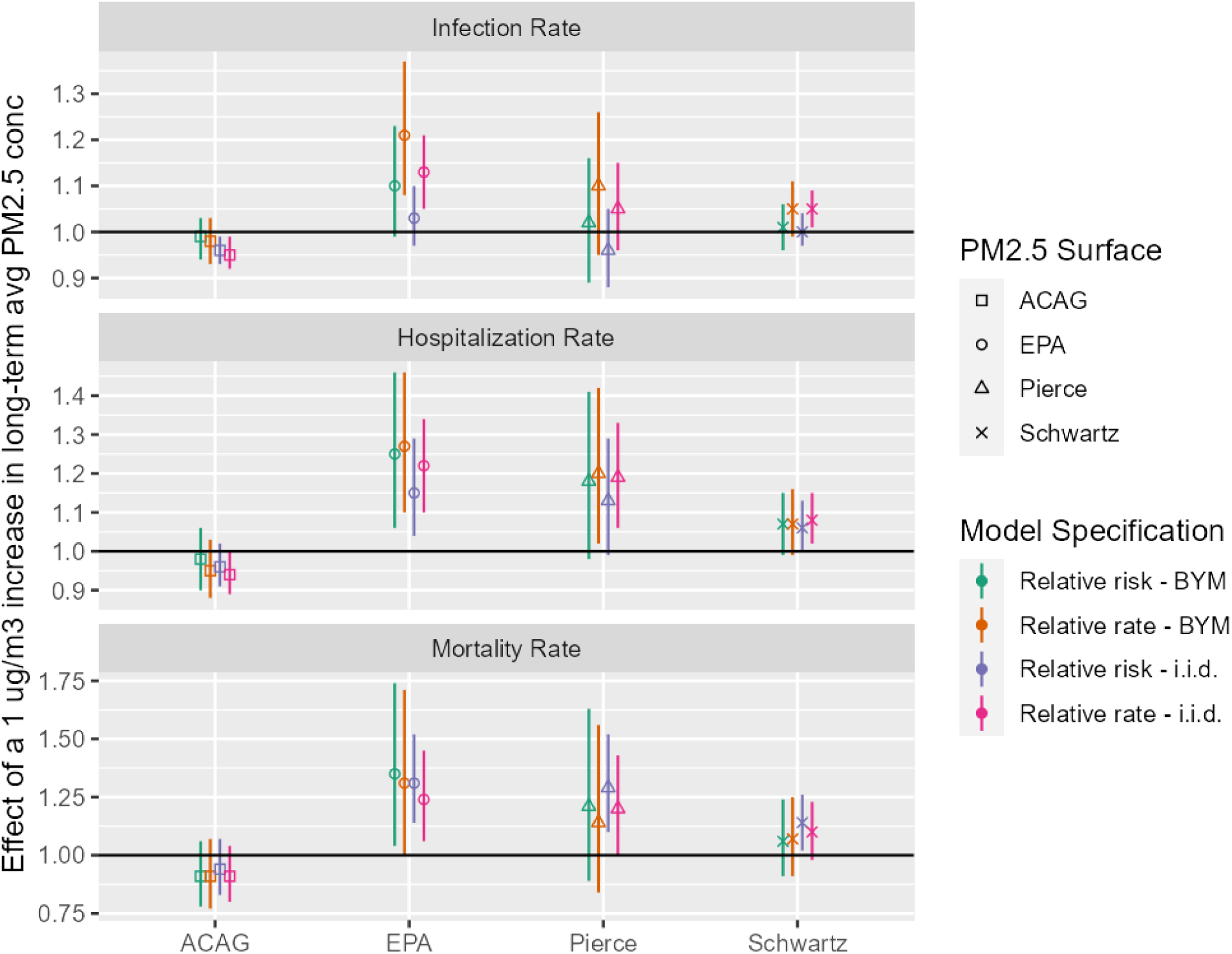
Estimated effect of a 1µg/m^3^ increase in long-term average PM2.5 concentration on three COVID outcomes, from fully adjusted models.

Table 2 displays DIC value and Moran’s I statistics from fully adjusted models for each of our four study designs using the EPA PM2.5 surface as the key exposure variable. DIC values and Moran’s I test results both indicated the relative risk - BYM design produced the best-fitting model. Given the improved fit from the relative risk-BYM model design and the robust results across all four designs, we focus primarily on this model for the remainder of this paper.

**Table 2:** Deviance Information Criteria (DIC) and Residual Moran’s I Statistic (p-value) by study design and outcome.

| Study Design | Infections | Hospitalizations | Mortality |
| --- | --- | --- | --- |
| Relative risk - BYM | 8542<br>-0.05 (p > 0.99) | 5179<br>-0.04 (p > 0.99) | 3042<br>-0.06 (p > 0.99) |
| Relative rate - BYM | 8654<br>-0.05 (p > 0.99) | 5212<br>-0.04 (p > 0.99) | 3064<br>-0.06 (p > 0.99) |
| Relative risk - i.i.d. | 8572<br>0.09 (p < .001) | 5183<br>0.02 (p = 0.12) | 3050<br>0.02 (p = 0.09) |
| Relative rate - i.i.d. | 8670<br>0.09 (p < .001) | 5218<br>0.02 (p = 0.12) | 3071<br>0.02 (p = 0.09) |

### PM2.5 effects

Our results clearly suggest the choice of exposure surface has a large impact on the direction and significance of the adjusted association between long-term PM2.5 exposure and COVID-19 outcomes. The EPA exposure surface displayed the most consistent positive adjusted associations with COVID-19 outcomes. After controlling for all other census-tract level factors, long-term PM2.5 exposure measured by the EPA surface was statistically associated with an increase in COVID-19 infections in two out of four study designs, and with increases in hospitalizations and mortality across all four study designs. According to our best-fitting model, a 1 µg/m^3^ increase in long-term PM2.5 exposure measured by the EPA surface was associated with a positive, but insignificant increase in the relative risk of COVID-19 infections (RR: 1.10, 95% CI: 0.99 - 1.21). The same increase in long-term PM2.5 exposure was associated with a 25% increase in the relative risk of hospitalizations (RR: 1.25, 95% CI: 1.06 - 1.46) and a 35% increase in the relative risk of mortality (RR: 1.35, 95% CI: 1.05 - 1.74). Results for the Pierce and Schwartz surfaces were generally consistent with those of the EPA surface; however, none of the associations for these two surfaces were statistically significant in the relative risk-BYM model. A 1 µg/m^3^ increase in long-term PM2.5 exposure measured by the Pierce surface was associated with positive, but insignificant increases in the relative risk of COVID-19 infections (RR: 1.02, 95% CI: 0.89 – 1.15), hospitalizations (RR: 1.18, 95% CI: 0.98 – 1.41), and mortality (RR: 1.21, 95% CI: 0.89 – 1.44). Likewise, the same increase in long-term PM2.5 exposure measured by the Schwartz surface was associated with positive, but insignificant increases in the relative risk of COVID-19 infections: (RR: 1.01, 95% CI: 0.96 - 1.06), hospitalizations (RR: 1.07, 95% CI: 0.99 - 1.15), and mortality (RR: 1.06, 95% CI: 0.91 - 1.24).

At the same time, the results from the ACAG surface point in the opposite direction from the three other exposure surfaces. In our best-fitting model, a 1 µg/m^3^ increase in long-term PM2.5 measured by the ACAG surface was associated with a statistically insignificant decrease in the relative risk of COVID-19 infections (RR: 0.95, 95% CI: 0.94 – 1.03), hospitalizations (RR: 0.98, 95% CI: 0.90 – 1.06), and mortality (RR: 0.91, 95% CI: 0.78 – 1.06).

### Other Effects

While not the primary goal of our analysis, we identified significant associations between several other factors and COVID-19 infections, hospitalizations, and mortality at the census - tract level in Colorado. Adjusting for all other census-tract-level factors, we found statistically significant evidence that census-tracts with a larger proportion of non-Hispanic African American population are associated with increased risk of COVID-19 infections (RR: 1.04, 95% CI: 1.02 - 1.06) and hospitalizations (RR: 1.07, 95% CI: 1.04 - 1.11). In addition, census-tracts with larger proportions of non-African American people of color were statistically significantly and positively associated with the risk of infections (RR: 1.31, 95% CI: 1.25 - 1.39), hospitalizations (RR: 1.44, 95% CI: 1.32 - 1.58), and mortality (RR: 1.59, 95% CI: 1.32 - 1.91). With respect to social distancing-related factors, we found a one-standard deviation increase in our mobility risk index was statistically associated with increases in the risk of infections (RR: 1.18, 95% CI: 1.09 - 1.28) and hospitalizations (RR: 1.15, 95% CI: 1.04 - 1.27).

In addition, we found evidence that communities with a larger proportion of essential workers were subject to increased risk of COVID-19 infections (RR: 1.05, 95% CI: 1.01 - 1.09) and hospitalizations (RR: 1.06, 95% CI: 1.00 - 1.13), whereas those with greater percentages of populations confined to correctional facilities, nursing homes, or mental hospitals had lower risk of hospitalizations (RR: 0.92, 95% CI: 0.87 - 0.97), but potentially higher risk of mortality (RR: 1.11, 95% CI:1.00 - 1.22). Despite indirect age adjustment of our estimates via an expected count in our offset term, we found statistically significant evidence that mortality risk is higher in both census tracts with greater population percentages of adults age 65 years and older (RR: 1.30, 95% CI: 1.14 - 1.49) and tracts with greater population percentages of adults age 18 to 44 years (RR: 1.37, 95% CI: 1.12 - 1.68). Lastly, the number of days since the first case in a census tract was diagnosed was positively associated with higher risk of infections (RR: 1.08, 95% CI: 1.04 - 1.11), hospitalizations (RR: 1.27, 95% CI: 1.16 - 1.40), and mortality (RR:1.33, 95% CI: 1.11 - 1.63), while the crude COVID-19 test rate was negatively associated with the risk of infection (RR: 0.96, 95% CI: 0.94 - 0.98) and hospitalization (RR: 0.91, 95% CI: 0.88 - 0.95). The relative risks and 95% CIs for all fixed effects in the fully adjusted relative risk-BYM model are shown in Fig. 5, for all 3 COVID-19 outcomes and all four PM2.5 surfaces.

**Figure 5:**
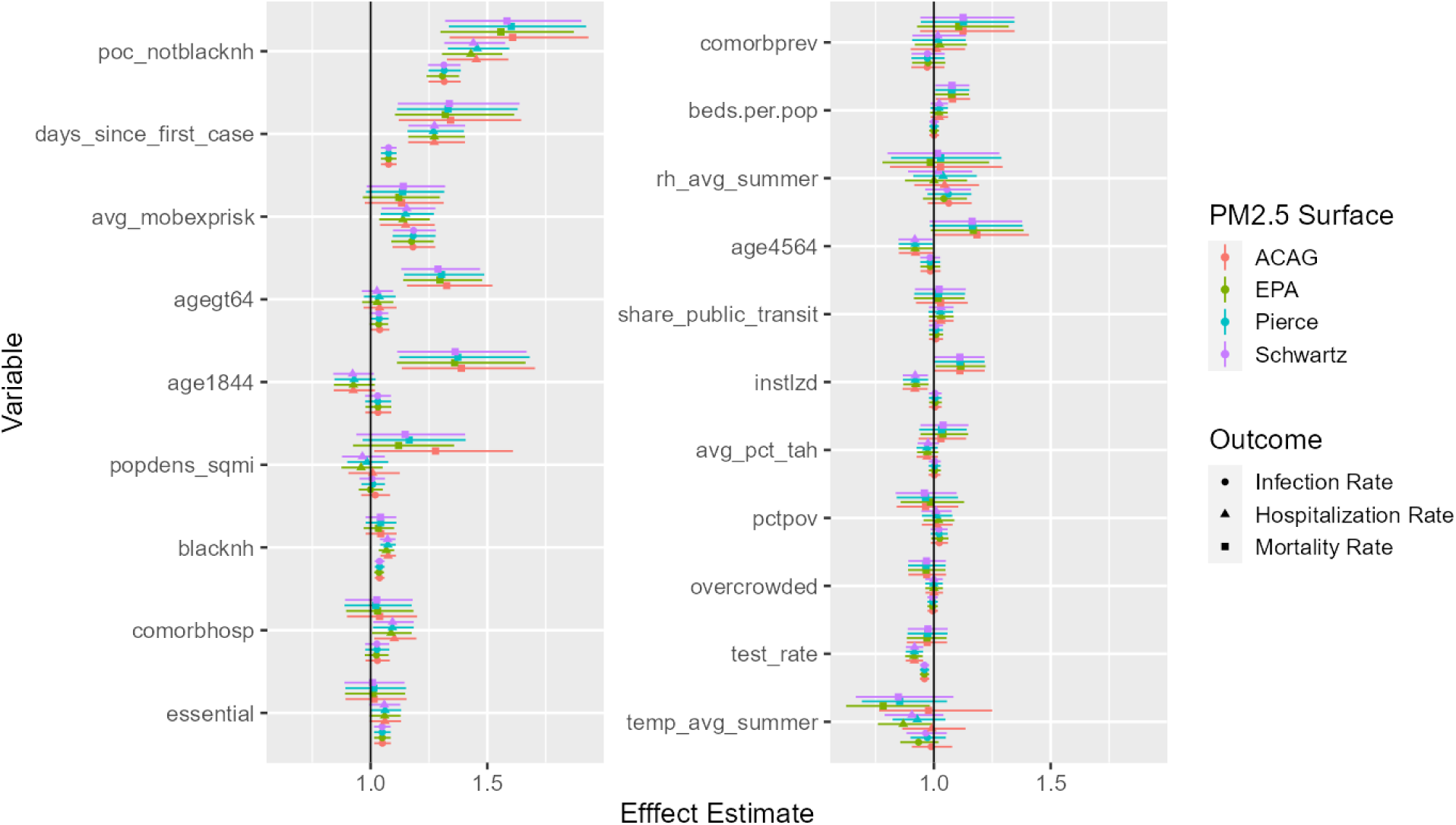
Effect estimates for all covariates included in the model. Points represent the estimated mean and thin lines represent the 95% confidence interval. Shapes indicate the COVID-19 outcome considered and colors the PM2.5 surface used.

## Discussion

### Long-term PM2.5 Exposure

Previous studies on the effects of long-term PM2.5 exposure on COVID-19 outcomes have reported a wide range of results. At least six studies have reported estimated effects for a 1 µg/m^3^ increase in PM2.5 concentrations on COVID-19 mortality at the county level in the United States (Knittel and Ozaltun, 2020; Wu et al., 2020; Tian et al., 2020; Liang et al., 2020; Rodriguez-Diaz et al., 2020; Millett et al., 2020). Estimates from these studies range from a non-significant 1% decrease (Knittel and Ozaltun, 2020) to a statistically significant 17% increase (Tian et al., 2020). The Liang et al., 2020 study used the Schwartz surface for their PM2.5 estimates, while the other five studies used the ACAG surface. In the United Kingdom, Travaglio et al., 2021 found that a 1 µg/m^3^ increase in PM2.5 can be associated with anywhere from a 4% decrease to a non-significant 1% increase, depending on how the data is aggregated temporally.

Previous studies looking at COVID-19 infections have reported even greater variability in the estimated PM2.5 effects. At the county level in the United States, Millet et al., 2020 and Rodriguez-Diaz et al., 2020 both report that a 1 µg/m^3^ increase in PM2.5 is associated with a non-significant 1% increase in COVID-19 infections. In the United Kingdom, Travaglio et al., 2021 find that estimated effect can range from a 10% decrease to a 12% increase, depending on how the data are aggregated; in the Netherlands, Andrée, 2020 found that PM2.5 from 8 µg/m^3^ to 10 µg/m^3^ is associated with a decrease in COVID-19 infections, while a further increase from 10 to 12 µg/m^3^ is associated with nearly a 100% increase in COVID-19 infections; In China, Zheng et al., 2020 report that a 1 µg/m^3^ increase in PM2.5 is associated with a statistically significant 1% increase in COVID-19 infections.

These studies used different model designs, covered different time periods, and adjusted for different covariates. It is therefore unsurprising that these studies found different estimates of the effect on PM2.5 on COVID-19 outcomes. Our analysis adds another potential source of uncertainty, that differences in how PM2.5 is estimated over small spatial regions can cause noticeable changes in the results. Among the 12 surface-outcome combinations studied here, results from our best-fitting model suggested 2 associations are positive and statistically significant, 7 are positive but not statistically significant, and 3 are negative but not statistically significant. None of our results from our best-fitting model in the statewide analysis or in the time-period or study area sensitivity studies show a statistically significant decrease in COVID-19 outcomes with increases in long-term PM2.5 concentrations.

To better understand which PM2.5 surfaces are most accurate in Colorado, Fig. 6 compares the different PM2.5 surfaces against EPA monitors in Colorado. The PM2.5 surface estimates for each census tract are compared against the average of all year-round PM2.5 monitors in that census tract, following the procedure described in Diao et al., 2019. Both federal reference method/federal equivalent method (FRM/FEM) monitors and non-FRM/FEM monitors were included. The EPA and Schwartz surfaces show the best agreement with the EPA monitors, with lower root mean squared error and mean absolute error than the other two surfaces.

**Figure 6:**
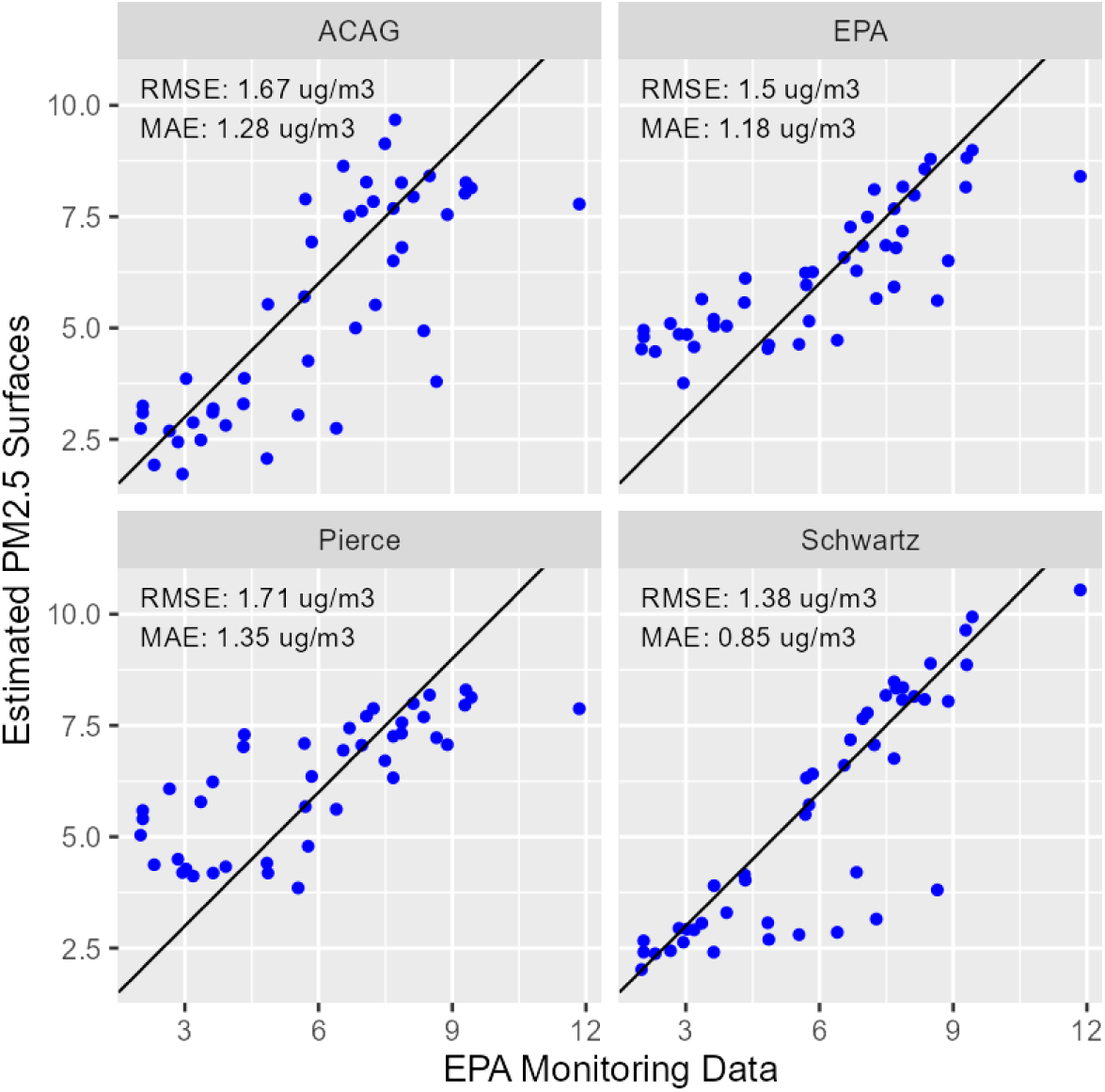
Comparison of PM2.5 surfaces with EPA monitors in Colorado. RMSE = root mean square error; MAE = mean absolute error.

However, the EPA monitors are not an independent comparison, since all four PM2.5 monitoring surfaces are built in part on the same EPA monitoring data. Furthermore, these comparisons cannot resolve the question of how the surfaces perform in areas far away from regulatory monitors. A more rigorous comparison of the performance of the PM2.5 surfaces would compare the PM2.5 surfaces against in-situ monitoring data that was not used in constructing the surface. Unfortunately, we were unable to locate any in-situ measurements other than the EPA monitoring data that reported year - round PM2.5 concentrations starting in 2016 or earlier. However, as these surfaces are updated for additional years, we are optimistic that non-regulatory monitors such as those used in the LoveMyAir network in Denver (DDPHE, 2020) or by PurpleAir (PurpleAir, 2020) will allow such an independent comparison to be made. Recent work to calibrate these monitors under a variety of conditions is another crucial step before these sensors can be used to check the performance of PM2.5 models (Holder et al., 2020; Considine et al., 2021). At the moment, without the ability to independently measure the error in the PM2.5 surfaces, it is impossible to identify which surfaces are most accurate in Colorado.

Looking forward, better estimates of the health effects of environmental exposures will require resolving the disagreements among the four PM2.5 estimation surfaces. Figure 7 shows where the surfaces currently agree or disagree, based on the standard deviation of the four surfaces in each census tract. The greatest areas of disagreement are typically found in the area around the Denver metro area, potentially suggesting that the disagreement is caused not by differences in PM2.5 emissions (which are likely to be concentrated in the metro core), but are caused by differences in the modeled chemical formation and transport of PM2.5.

**Figure 7:**
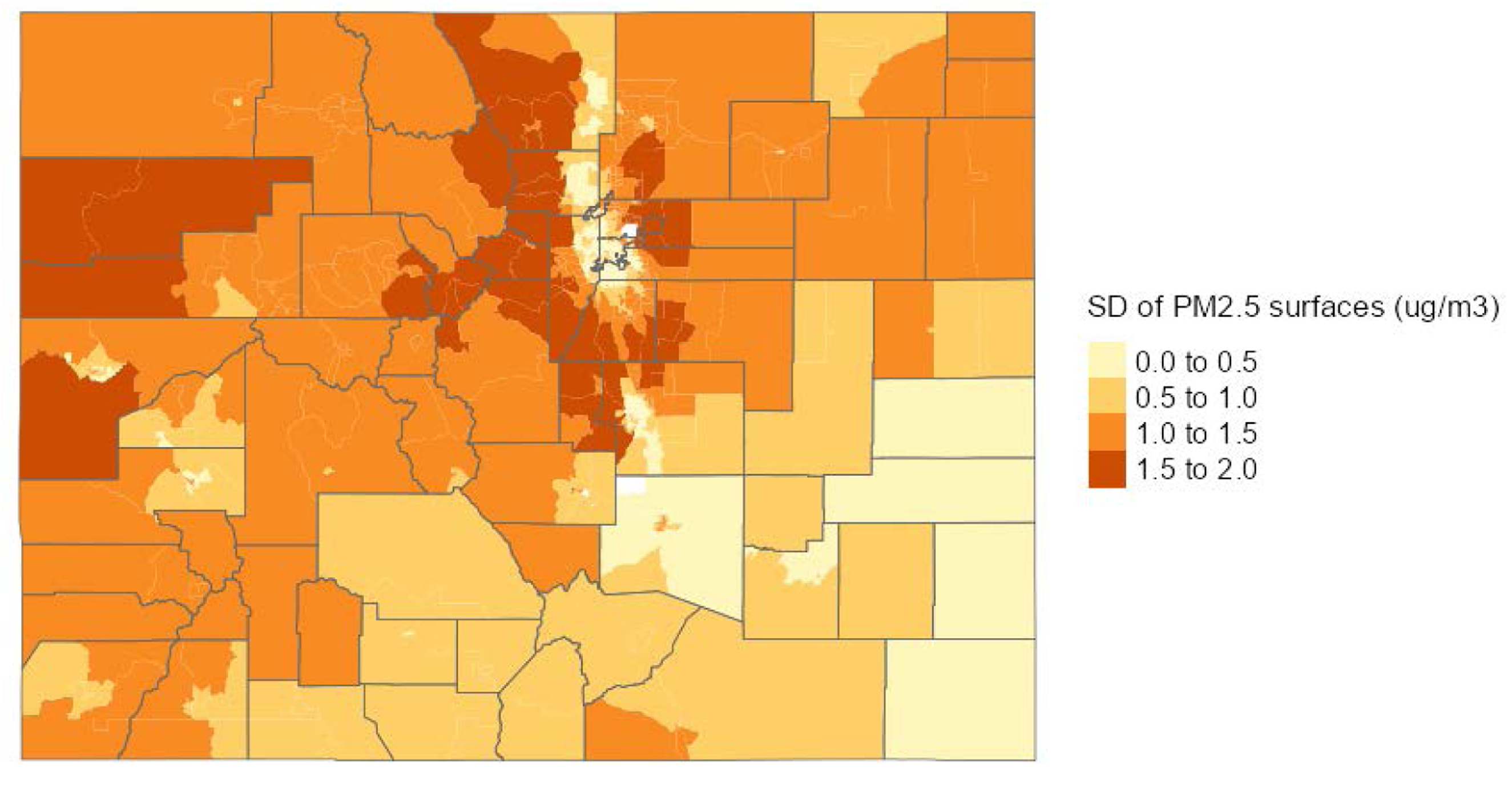
Standard deviation of the four PM2.5 estimates in each census tract

Simply examining where the PM2.5 surfaces agree or disagree does not tell us which of the census tracts have the greatest impact on our model results. We therefore examined how our regression results changed if we changed the PM2.5 estimate in a single census tract. Figure 8 shows the change in our PM2.5 effect estimate on hospitalizations when the PM2.5 estimate for a single census tract is changed from the EPA surface to the ACAG surface. The EPA and ACAG surfaces we used in this sensitivity analysis as these surfaces showed the highest and lowest PM2.5 effects (Fig. 4).

**Figure 8:**
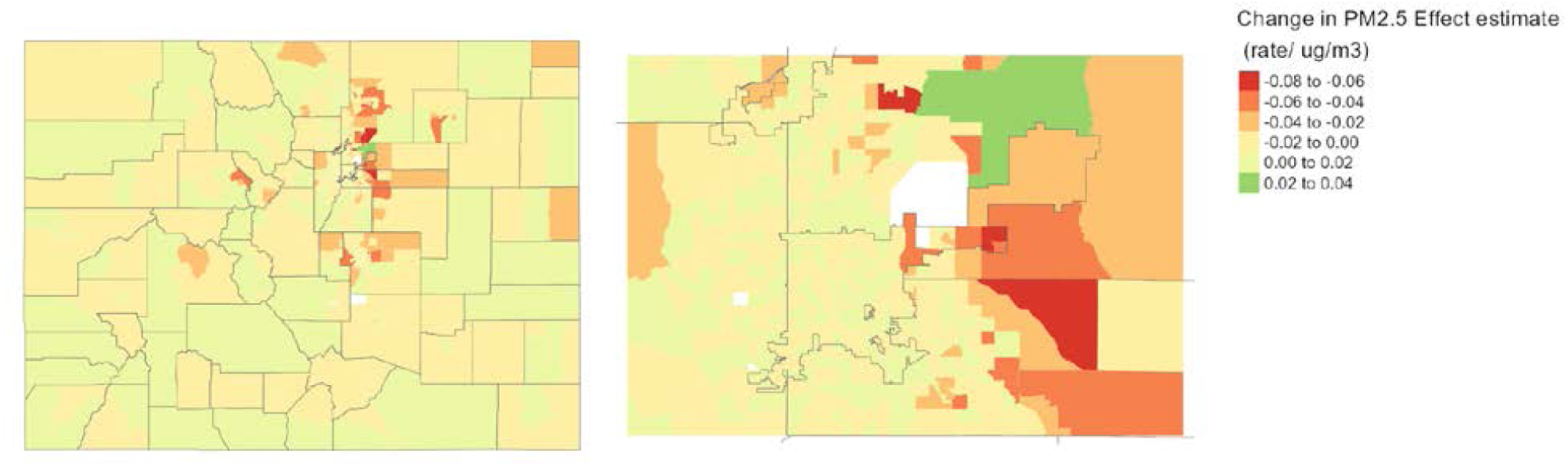
Change in PM2.5 effect estimate when the EPA PM2.5 estimates are changed to the ACAG estimates.

The change in PM2.5 coefficient is generally negative, which matches the large decrease in PM2.5 effect estimate when the PM2.5 estimate in all census tracts is changed from the EPA surface to the ACAG surface. The largest change in PM2.5 coefficients are generally found when the PM2.5 surfaces are changed in census tracts running along the eastern edge of the Front Range Urban Corridor. This region combined high disagreement between PM2.5 surfaces, high numbers of COVID-19 hospitalizations, and relatively high population. The combination of these three factors means that changes in the PM2.5 estimate in this region have relatively large effects on our overall results. The high leverage of this region suggests that improving the accuracy of our PM2.5 estimates in this area could significantly improve our ability to link environmental exposures to health outcomes in Colorado.

### High Risk Populations

Early descriptions of outcomes among patients with COVID-19 in the United States indicated disease severity is highest among adults age 65 years and older, people with underlying health conditions, and is strongly linked to race/ethnicity and other socioeconomic factors (CDC COVID-19 Response Team, 2020a, 2020b; Hsu et al. 2020). Health inequities of the COVID-19 pandemic are thought to be at least partially explained by well-documented racial and ethnic disparities in the prevalence of chronic health conditions (Alcendor, 2020); however, several recent studies have suggested disparities among Black and Hispanic/Latino communities may be better explained by overrepresentation among the essential workforce (Oppel et al. 2020; Rogers et. al. 2020). While our ecological study designs means we cannot make inferences about individuals, our results offer a proxy for understanding how these patterns increase vulnerability disproportionately for certain populations in Colorado.

Our findings suggest communities of color in Colorado are subject to higher risk of infection as well as of more severe complications of COVID-19. Using the fully adjusted relative risk-BYM model we found statistically significant associations between larger proportions of non-Hispanic African Americans and increased risk of COVID-19 infections and hospitalizations regardless of the choice of PM2.5 surface, while the association with the risk of COVID-19 mortality was positive but not statistically significant. We also found that larger proportions of non-African American people of color, a group largely composed of Colorado’s Hispanic/Latino community, were statistically associated with increased risk of all three COVID-19 outcomes. Further, we found a separate statistically significant association between the percentage of census-tract population employed in an essential industry and the risk of COVID-19 infection, which appears to be further supported by our finding that communities with more individuals in prime working age (18-44 years) are subject to greater risk of mortality.

Our findings are consistent with Millet et al., 2020 who in a similar ecological study of U.S. counties found that higher proportions of non-Hispanic African American residents were associated with increased rates of infection and mortality. With respect to non-Hispanic African Americans, the similarity of our findings to those of a nationwide study are particularly noteworthy given Colorado’s smaller overall population of non-Hispanic African Americans compared to other regions of the United States. A per-IQR increase in the proportion of non-Hispanic African Americans among Colorado census-tracts represents an increase of only 3.3% (0.3%, 3.6%), while an identical increase among all U.S. counties represents a 9.6% increase (0.7%, 10.3%). By contrast, a similar ecological study of U.S. counties by Rodriguez-Diaz et al., 2020 did not find statistically significant associations between predominantly Hispanic/Latino counties and COVID-19 rates in the Western U.S. Our findings suggest sub-county variation in the population of people of color may be crucial to understanding health disparities of the COVID-19 pandemic in Colorado. Together, the results of our study contribute additional support for the need to address the disproportionate impact of COVID-19 on higher-risk populations as the pandemic response and recovery efforts continue in Colorado.

## Conclusion

This study quantifies the environmental, demographic, and socioeconomic associations and disparities for three COVID-19 outcomes in Colorado. We show the uncertainty in census-tract PM2.5 concentrations is a key factor limiting our ability to link environmental exposures and health outcomes, and highlight specific regions in Colorado where the uncertainty in PM2.5 concentrations is most impactful.

In the majority of our analyses, we find that a small increase in long-term exposure to fine particle pollution is associated with a positive but non-statistically significant increase in infections, hospitalizations, and deaths due to COVID-19. It is notable that we see these results even at relatively low concentrations of PM2.5. The average concentration of PM2.5 in Colorado ranges from 1.78 and 9.95 µg/m^3^, below the World Health Organization’s guideline value for PM2.5 and meeting the EPA’s annual National Ambient Air Quality Standard. Our findings are consistent with studies showing a consistent effect of PM2.5 on daily mortality with no evidence of a threshold (Liu et al., 2019)

However, we are limited in the conclusions we can draw by the uncertainty in PM2.5 concentrations at the census-tract level. In the short-term, other researchers examining the links between PM2.5 and COVID-19 should carefully consider how uncertainty in PM2.5 concentrations affects their results. This is especially true when attempting to understand risk factors in Colorado over small spatial scales or in neighborhoods with dramatically different disease rates.

In the long-term, improved understanding of the links between air pollution and human health at the neighborhood scale will require the development of improved PM2.5 surfaces that can more accurately capture variations in PM2.5 concentrations on the scale of kilometers. Increased monitoring, either from regulatory or low-cost sensors, higher resolution computer models, and remote sensing products from new satellites could all play a role in improving the accuracy and precision of PM2.5 surfaces at the neighborhood scale. In addition, there is a clear need for a method to independently validate these PM2.5 surfaces after they are generated.

While the uncertainty in PM2.5 concentrations makes it difficult to establish definitive conclusions, this study provides evidence that local rates of COVID-19 infections, hospitalizations, and mortality are influenced by patterns of exposure to air pollution, racial and ethnic composition, local travel patterns, and risk factors such as aging population structure and underlying comorbidities. As such, this study could help provide a scientific basis for precisely targeting COVID-19 response efforts in Colorado based upon community-specific risk factors. For example, additional public health interventions, such as recommending ways to decrease personal exposure to fine particle pollution and considering ways to effect more social distancing, could be implemented in specific neighborhoods to better protect those most at risk.

Finally, the results of this analysis indicate the need for expanded investigations regarding the link between air pollution exposure and COVID-19 outcomes. In particular, further analysis of the link between short-term air pollution exposure and COVID-19 is urgently needed to establish whether these more modifiable exposures play a role in infection and disease severity. The impact of long-term PM2.5 exposure on COVID-19 outcomes at relatively low levels of PM2.5 pollution is another area that would be helped by further research.

## Data Availability

COVID-19 outcomes at the census-tract level cannot be released publically; however, other datasets are publicly available. All code to complete this analysis is available upon request to the project authors.

## Acknowledgements

We thank Dr. Colleen Reid (University of Colorado Boulder); Dr. Jennifer Peel, Dr. Andreas Neophytou, and Dr. Sheryl Magzamen (Colorado State University); and Dr. John Adgate and Dr. Jonathan Samet (University of Colorado Anschutz) for their critical review of an early draft manuscript. We thank Xiao Wu, Benjamin Sabath, and Dr. Francesca Dominici (Harvard University) for their valuable feedback on our regression model. We thank Dr. Joel Schwartz (Harvard University) for providing access to modeled PM2.5 data for Colorado. We also thank Shannon Barbare (Colorado Department of Public Health & Environment) for proofreading the final manuscript.

This work was supported by the Grant or Cooperative Agreement Number, 5 NUE1EH001346-04-00, funded by the Centers for Disease Control and Prevention. Its contents are solely the responsibility of the authors and do not necessarily represent the official views of the Centers for Disease Control and Prevention or the Department of Health and Human Services.

## Statement on Structural Inequity

The Colorado Department of Public Health and Environment acknowledges that long-standing systemic racism, including economic and environmental injustice, has created conditions that negatively affect marginalized communities, particularly people of color. These conditions, which limit opportunities for optimal health and influence individual behaviors, are critical predictors of health outcomes. To realize a future where all Coloradans can thrive, we must be leaders in undoing policies and practices that have contributed to these inequities.

