## Supporting Information for "Long-term air pollution and other risk factors associated with COVID-19 at the census-tract-level in Colorado"

#### Population Density in Colorado

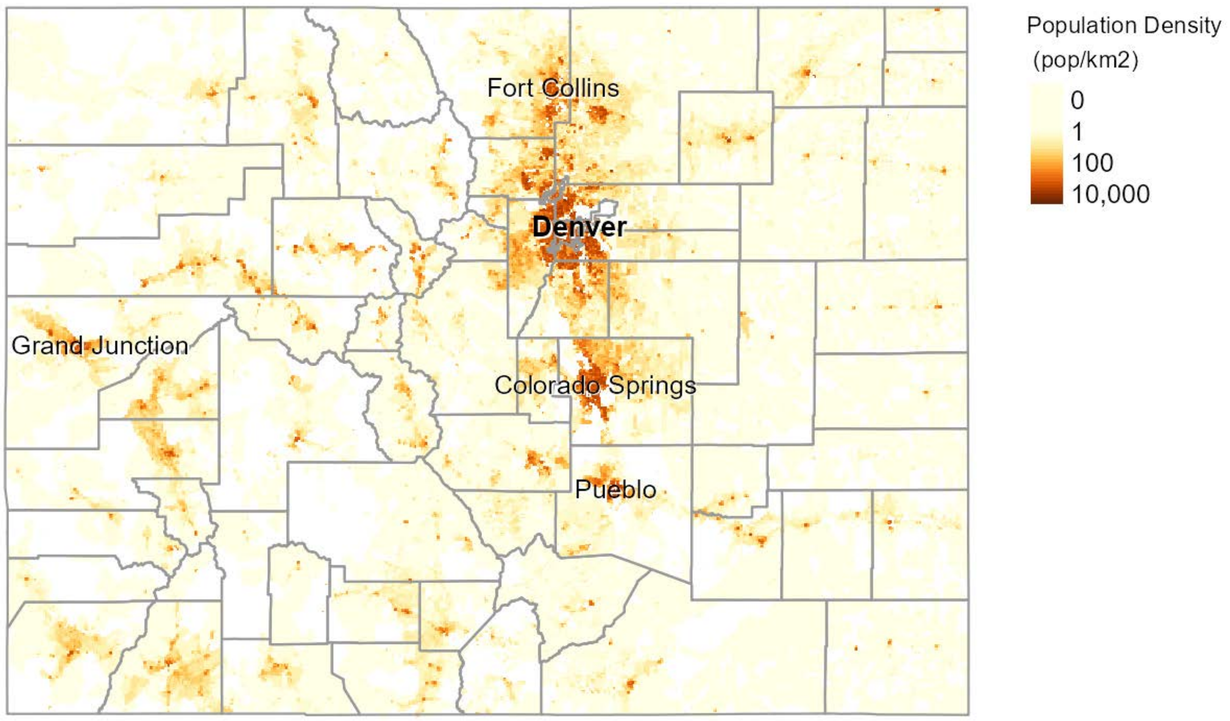


Figure S1: Population density and major cities in Colorado, based on the 2010 Census

#### Variable descriptions

| Variable Name | Description | Data Source |
|---|---|---|
| age1844 | The percent of census tract population ages 18 to 44 years. | U.S. Census Bureau, American Community Survey 5-year Estimates (2014-2018). Table: B01001 |
| age4564 | The percent of census tract population ages 45 to 64 years. | U.S. Census Bureau, American Community Survey 5-year Estimates (2014-2018). Table: B01001 |
| agegt64 | The percent of census tract population age 65 years and older. | U.S. Census Bureau, American Community Survey 5-year Estimates (2014-2018). Table: B01001 |

| | | |
|---|---|---|
| asthma_hosp | Age-adjusted Colorado census tract rate of asthma-related hospital discharges (2013-2017) per 100,000 persons. The rates are calculated using the geocoded billing address of discharged individuals found in the dataset with the selected icd-10 codes and 2013-2017 population estimates from the American Community Survey. | Colorado Hospital Association. https://data-cdphe.opendata.arcgis.com/datasets/cdphe-composite-selected-health-outcome-dataset-census-tract |
| avg_mobexprisk | Derived measure of SARS-COV2 exposure that combines daily travel to a census tract with the case rate of the corresponding county. Higher measures correspond to more travel originating in counties with higher COVID-19 case rates. | Mobile phone data source: Safegraph COVID-19 Data Consortium https://www.safegraph.com/covid-19-data-consortium<br>Data prepared by Citizen Software Engineers. |
| avg_no2_ppb | Long-term annual average (2000-2010) Nitrogen Dioxide concentration in parts per billion (ppb) | National Spatiotemporal Exposure Surface for NO2: Monthly Scaling of a Satellite-Derived Land-Use Regression, 2000-2010. Matthew J. Bechle, Dylan B. Millet, and Julian D. Marshall. Environmental Science & Technology 2015 49 (20), 12297-12305. DOI: 10.1021/acs.est.5b02882. |
| avg_o3_ppb | Long-term annual average (2002-2016) ozone concentration in parts per billion (ppb) | U.S. EPA Fused Air Quality Surface Using Downscaling (FAQSD) files. https://www.epa.gov/hesc/rsig-related-downloadable-data-files |
| avg_pct_tah | Measure of the average daily amount of time spent at home per census tract derived from mobile phone location data | Mobile phone data source: Safegraph COVID-19 Data Consortium https://www.safegraph.com/covid-19-data-consortium<br>Data prepared by Citizen Software Engineers. |
| beds.per.pop | Number of hospital beds per population in each census tract. Population Estimates from the American Community Survey. | Homeland Infrastructure Foundation-Level Data (HIFLD). https://hifld-geoplatform.opendata.arcgis.com/datasets/hospitals |

| | | |
|---|---|---|
| blacknh | Percent of census tract population that is Black or African American (Non-Hispanic). | U.S. Census Bureau, American Community Survey 5-year Estimates (2014-2018). Table: B01001B |
| comorbhosp | Composite measure created via Principal Component Analysis. Strongly correlated with age-adjusted rate of hospitalization for asthma, diabetes, heart disease, and influenza. | Colorado Hospital Association. [https://data-cdphe.opendata.arcgis.com/datasets/cdphe-composite-selected-health-outcome-dataset-census-tract](https://data-cdphe.opendata.arcgis.com/datasets/cdphe-composite-selected-health-outcome-dataset-census-tract) |
| comorbprev | Composite measure created via Principal Component Analysis. Strongly correlated with prevalence of asthma, diabetes, heart disease, obesity, smoking, and poor health status. | Colorado Department of Public Health & Environment. [https://data-cdphe.opendata.arcgis.com/search?q=community%20level%20estimates](https://data-cdphe.opendata.arcgis.com/search?q=community%20level%20estimates) |
| days_since_first_case | Number of days since the first COVID-19 case was reported for a census tract. | Colorado Department of Public Health & Environment. |
| diabetes | These data represent the predicted (modeled) prevalence of diabetes among adults (Age 18+) for each census tract in Colorado. | Colorado Department of Public Health & Environment. [https://data-cdphe.opendata.arcgis.com/search?q=community%20level%20estimates](https://data-cdphe.opendata.arcgis.com/search?q=community%20level%20estimates) |
| diabetes_hosp | Age-adjusted Colorado census tract rate of diabetes-related hospital discharges (2013-2017) per 100,000 persons. The rates are calculated using the geocoded billing address of discharged individuals found in the dataset with the selected icd-10 codes and 2013-2017 population estimates from the American Community Survey. | Colorado Hospital Association. [https://data-cdphe.opendata.arcgis.com/datasets/cdphe-composite-selected-health-outcome-dataset-census-tract](https://data-cdphe.opendata.arcgis.com/datasets/cdphe-composite-selected-health-outcome-dataset-census-tract) |
| eduhths | Percent of census tract adult population (18+) that did not graduate from high school or obtain a GED. | U.S. Census Bureau, American Community Survey 5-year Estimates (2014-2018). Table: B06009 |
| egltvw.esp | Percent of census tract population whose primary language is Spanish and who | U.S. Census Bureau, American Community Survey 5-year Estimates (2014-2018). Table: B06007 |

| | | |
|---|---|---|
| | speak English less than very well. | |
| essential | This is the percentage of the population working in an essential sector. The essential workforce was determined by Colorado's stay-at-home order and includes those working in 10 occupational codes. Data include those working in agriculture, construction, manufacturing, wholesale trade, retail trade, transportation, waste management, health care and social assistance, food services, and other services such as child care and banks. | U.S. Census Bureau, American Community Survey 5-year Estimates (2014-2018). Table: S2404 |
| flu_hosp | Age-adjusted Colorado census tract rate of influenza-related hospital discharges (2013-2017) per 100,000 persons. The rates are calculated using the geocoded billing address of discharged individuals found in the dataset with the selected icd-10 codes and 2013-2017 population estimates from the American Community Survey. | Colorado Hospital Association. https://data-cdphe.opendata.arcgis.com/datasets/cdphe-composite-selected-health-outcome-dataset-census-tract |
| hd_hosp | Age-adjusted Colorado census tract rate of heart disease-related hospital discharges (2013-2017) per 100,000 persons. The rates are calculated using the geocoded billing address of discharged individuals found in the dataset with the selected icd-10 codes and 2013-2017 population estimates from the American Community Survey. | Colorado Hospital Association. https://data-cdphe.opendata.arcgis.com/datasets/cdphe-composite-selected-health-outcome-dataset-census-tract |
| heartdisease | These data represent the predicted (modeled) prevalence of heart disease among adults (Age 18+) for each census tract in Colorado. | Colorado Department of Public Health & Environment. https://data-cdphe.opendata.arcgis.com/search?q=community%20level%20estimates |

| | | |
|---|---|---|
| hispanic | Percent of census tract population that is Hispanic or Latino. | U.S. Census Bureau, American Community Survey 5-year Estimates (2014-2018). Table: B01001I |
| instlzd | Percent of population living in correctional facilities, nursing homes, or mental hospitals. | U.S. Census Bureau, American Community Survey 5-year Estimates (2014-2018). Table: S2603 |
| insured | Percent of census tract population with health insurance. | U.S. Census Bureau, American Community Survey 5-year Estimates (2014-2018). Table: S2704 |
| medhhinc | Median household income of a census tract. | U.S. Census Bureau, American Community Survey 5-year Estimates (2014-2018). Table: B25077 |
| obesity | These data represent the predicted (modeled) prevalence of obesity among adults (Age 18+) for each census tract in Colorado. | Colorado Department of Public Health & Environment. https://data-cdphe.opendata.arcgis.com/search?q=community%20level%20estimates |
| overcrowded | Percent of occupied housing units with more than 1 person per bedroom. | U.S. Census Bureau, American Community Survey 5-year Estimates (2014-2018). Table: B25014 |
| ownerocc | The percent of census tract occupied housing units that are owner-occupied. | U.S. Census Bureau, American Community Survey 5-year Estimates (2014-2018). Table: B25008 |
| pctpov | The percent of census tract population living in households with family income in the last 12 months below the federal poverty line. | U.S. Census Bureau, American Community Survey 5-year Estimates (2014-2018). Table: B06012. |
| poc_notblacknh | Percent of census tract population that is neither African American nor White (Non-Hispanic). | U.S. Census Bureau, American Community Survey 5-year Estimates (2014-2018). |
| popdens_sqmi | Population per square mile in each census tract. | U.S. Census Bureau, American Community Survey 5-year Estimates (2014-2018). Table: B01001 |
| rh_avg_summer | 30-year normal average summer relative humidity in each census tract. | PRISM Climate Group, Oregon State University, http://prism.oregonstate.edu, created 1 June 2020. |

| | | |
|---|---|---|
| rh_avg_winter | 30-year normal average winter relative humidity in each census tract. | PRISM Climate Group, Oregon State University, http://prism.oregonstate.edu, created 1 June 2020. |
| share_bike | The percent of census tract population (16+) who commute to work by bicycle. | U.S. Census Bureau, American Community Survey 5-year Estimates (2014-2018). Table: B08301 |
| share_drive | The percent of census tract population (16+) who commute to work by car, truck, or van. | U.S. Census Bureau, American Community Survey 5-year Estimates (2014-2018). Table: B08301 |
| share_public_transit | The percent of census tract population (16+) who commute to work by public transportation. | U.S. Census Bureau, American Community Survey 5-year Estimates (2014-2018). Table: B08301 |
| share_walk | The percent of census tract population (16+) who commute to work by walking. | U.S. Census Bureau, American Community Survey 5-year Estimates (2014-2018). Table: B08301 |
| smoking | These data represent the predicted (modeled) prevalence of smoking among adults (Age 18+) for each census tract in Colorado. | Colorado Department of Public Health & Environment. https://data-cdphe.opendata.arcgis.com/search?q=community%20level%20estimates |
| temp_avg_summer | 30-year normal average summer temperature in each census tract. | PRISM Climate Group, Oregon State University, http://prism.oregonstate.edu, created 1 June 2020. |
| temp_avg_winter | 30-year normal average winter temperature in each census tract. | PRISM Climate Group, Oregon State University, http://prism.oregonstate.edu, created 1 June 2020. |
| test_rate | Total unique individuals with a COVID-19 test divided by census tract population. Population estimates from the American Community Survey. | Colorado Department of Public Health & Environment. |
| whitenh | Percent of census tract population that is White (Non-Hispanic). | U.S. Census Bureau, American Community Survey 5-year Estimates (2014-2018). Table: B01001A |

Table S1: All potential confounding variables considered during the stepwise variable selection, definitions, and data sources.

#### Variable Selection Procedure

The reduced set of covariates was selected from the universe of 42 potential covariates through stepwise regression and manual examination of the variance inflation factors (VIFs). For speed and ease of use, we applied stepwise regression to a Poisson model using the EPA PM2.5 surface as our PM2.5 covariate. To account for the spatial co-linearity within the generalized linear model framework, spatial eigenvectors were included as additional covariates using the spmoran package (Murakami, 2019). The stepAIC function from the MASS package (Venables and Ripley, 2002) was then used to select an improved model, as measured by the Akaike information criterion (AIC). The variables selected during the stepwise regression procedure for COVID-19 infections are presented in the first column of Table S2.

We then further removed variables to reduce multicollinearity among the covariates, as measured by VIFs. Higher VIFs indicate additional multicollinearity, and a VIF of 10 or more is typically used to indicate severe multicollinearity (Kennedy, 1998). We removed an additional 21 covariates to reduce multicollinearity, either because those covariates had VIFs over 10, or because they were combined into other covariates (e.g., comorbhosp includes contributions from asthma_hosp, diabetes_hosp, hd_hosp, and flu_hosp). The selected variables after VIF filtering for the infection model are shown in the second column of Table S2.

Finally, we repeated this procedure for each of the additional two COVID-19 outcomes (hospitalizations and deaths). We then constructed a unified set of covariates that included all the covariates selected for any of the three outcomes. Adding these additional variables to the simplest models for each outcome did not greatly increase the VIF values for any of the covariates. The selected variables and their VIF values for the unified model are shown in the right-most column of Table S2.

| | **VIFs** | | |
|---|---|---|---|
| **Variable** | **After stepwise selection** | **After VIF filtering** | **Unified** |
| avg_pm25_ugm3 | 9.0 | 3.7 | 4.1 |
| avg_mobexprisk | 2.6 | 1.3 | 1.7 |
| comorbhosp | 25.5 | 2.2 | 2.3 |
| blacknh | 2.2 | 1.4 | 1.6 |
| pctpov | 4.4 | 2.4 | 2.7 |
| popdens_sqmi | 4.0 | 2.7 | 2.9 |
| essential | 3.5 | 2.2 | 2.3 |
| instlzd | 1.3 | 1.1 | 1.3 |
| age1844 | 8.5 | 5.0 | 5.2 |

| | | | |
|---|---|---|---|
| age4564 | 4.1 | 3.7 | 3.7 |
| agegt64 | 4.7 | 2.4 | 2.5 |
| temp_avg_summer | 31 | 2.7 | 3.4 |
| test_rate | 1.9 | 1.4 | 1.4 |
| days_since_first_case | 1.3 | 1.2 | 1.2 |
| share_public_transit | 4.0 | 1.6 | 1.8 |
| poc_notblacknh | . | 3.4 | 4.0 |
| avg_pct_tah | . | . | 1.6 |
| comorbprev | . | . | 2.8 |
| overcrowded | . | . | 1.4 |
| rh_avg_summer | . | . | 1.7 |
| beds.per.pop | . | . | 1.0 |
| avg_o3_ppb | 12 | . | . |
| avg_no2_ppb | 14 | . | . |
| hispanic | 12 | . | . |
| medhhinc | 7.8 | . | . |
| temp_avg_winter | 47 | . | . |
| rh_avg_winter | 10 | . | . |
| obesity | 12 | . | . |
| smoking | 3.2 | . | . |
| diabetes | 6.3 | . | . |
| heartdisease | 4.1 | . | . |
| asthma_hosp | 5.6 | . | . |
| diabetes_hosp | 5.2 | . | . |
| hd_hosp | 2.7 | . | . |
| flu_hosp | 3.2 | . | . |
| insured | 6.7 | . | . |
| ownerocc | 4.5 | . | . |
| edulths | 11 | . | . |
| engltvw.esp | 7.6 | . | . |
| share_drive | 10 | . | . |
| share_bike | 2.8 | . | . |
| share_walk | 4.6 | . | . |

Table S2: VIF analysis and stepwise selection results for confounding variables

### Study-area sensitivity analysis

According to our study area sensitivity analysis, there were statistically significant associations between PM2.5 exposure and all three COVID-19 outcomes among census tracts outside of the five most populous counties in the Denver metropolitan area (Figure S2). We found evidence that a 1 $\mu g/m^3$

increase in long-term PM2.5 exposure measured by the EPA surface was associated with increases in the relative risk of infections (RR: 1.19, 95% CI: 1.01 - 1.40), hospitalizations (RR: 1.45, 95% CI: 1.15 - 1.81), and mortality (RR: 1.54, 95% CI: 1.02 - 2.08). Further, posterior means for the Pierce and Schwartz surfaces were both greater than 1.0 in the models for hospitalizations and mortality, but their credible intervals included the null value. Meanwhile, none of the associations between any of the four PM2.5 exposure surfaces and COVID-19 outcomes were statistically significant among census tracts within the five most populous counties in the Denver metropolitan area. This finding was somewhat unexpected, although it may be explained by the relative homogeneity of PM2.5 exposure within the Denver metropolitan area as measured by any of the four exposure surfaces.

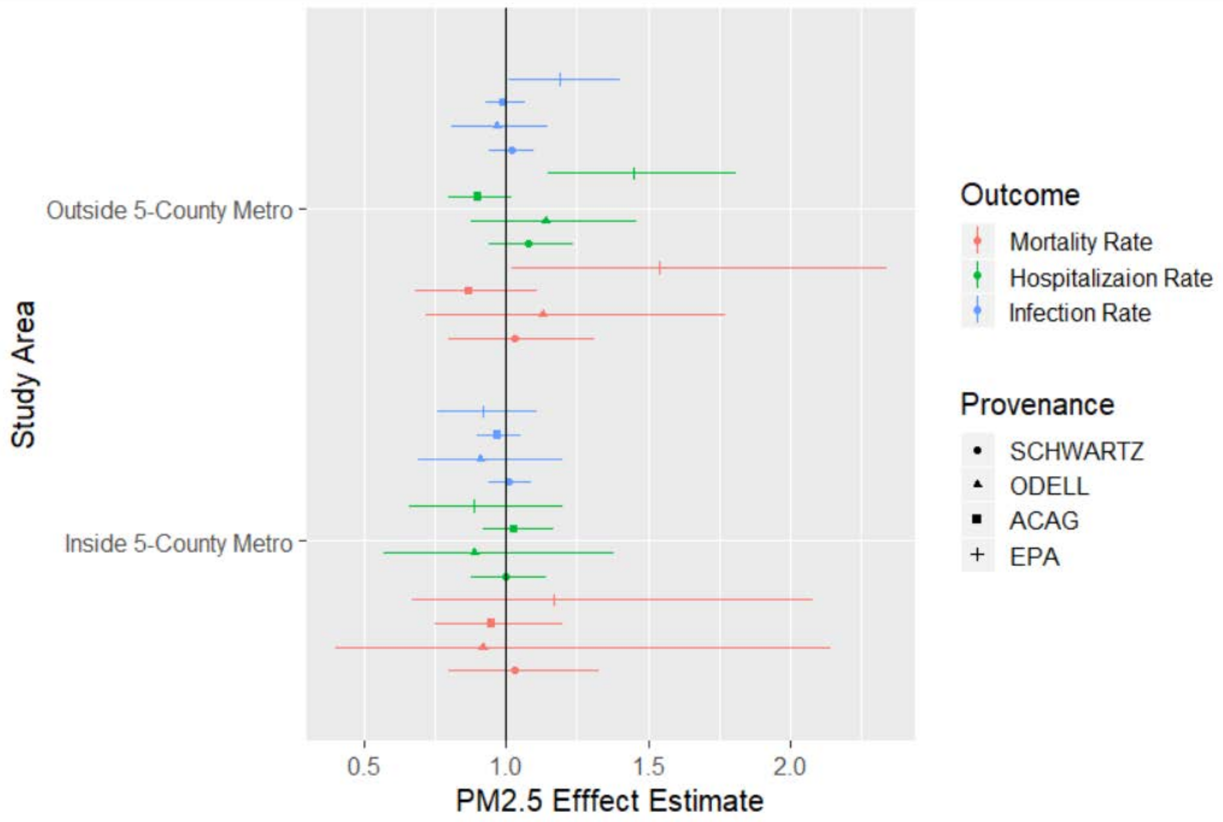


Figure S2: PM2.5 effect estimate outside and inside of the 5-county metro area.

#### Time period sensitivity analysis

In the time period sensitivity analysis we found the strongest evidence for a statistical association between long-term PM2.5 exposure and COVID-19 hospitalizations. From March 1, to May 9, 2020, we found statistically significant associations for the EPA (RR: 1.15, 95% CI: 1.03 - 1.30) and Pierce surfaces (RR: 1.16, 95% CI: 1.02 - 1.33), while the Schwartz surface suggested the association was positive, but insignificant (RR: 1.06, 95% CI: 0.99-1.15). Moreover, over the two subsequent time periods, posterior means for all four PM2.5 surfaces were greater than the null value. From May 9 to June 30, we observed statistically significant associations for the EPA (RR: 1.25, 95% CI: 1.05 - 1.54) and Schwartz surfaces (RR: 1.13, 95% CI: 1.01 - 1.28), but both the Pierce (RR: 1.19, 95% CI: 1.00 - 1.46) and ACAG (RR: 1.08, 95% CI: 0.96 - 1.23) surfaces suggested the association was positive but not significant. From July 1 to August 31, we observed statistically significant associations for the EPA (RR: 1.26, 95% CI: 1.07 - 1.52) and Pierce (RR: 1.25, 95% CI: 1.06 - 1.52) surfaces, while the ACAG (RR: 1.01, 95% CI: 0.92 - 1.12) and Schwartz (RR: 1.11, 95% CI: 1.00 - 1.23) surfaces reflected positive but insignificant associations. With respect to COVID-19 infections, our best fitting model produced posterior means for all four PM2.5 surfaces that were greater than the null value during each time period, and this association was positive and significant for all four PM2.5 surfaces during the two latter time periods. Lastly, with respect to COVID-19 mortality, the results were generally consistent with main analysis during all three time periods; however, the only statistically significant associations were found for the EPA (RR: 1.25, 95% CI: 1.02 - 1.59) surface from March 1 to May 9, and the Schwartz (RR: 1.32 95% CI: 1.01 - 1.78) surface from July 1 to August 31.

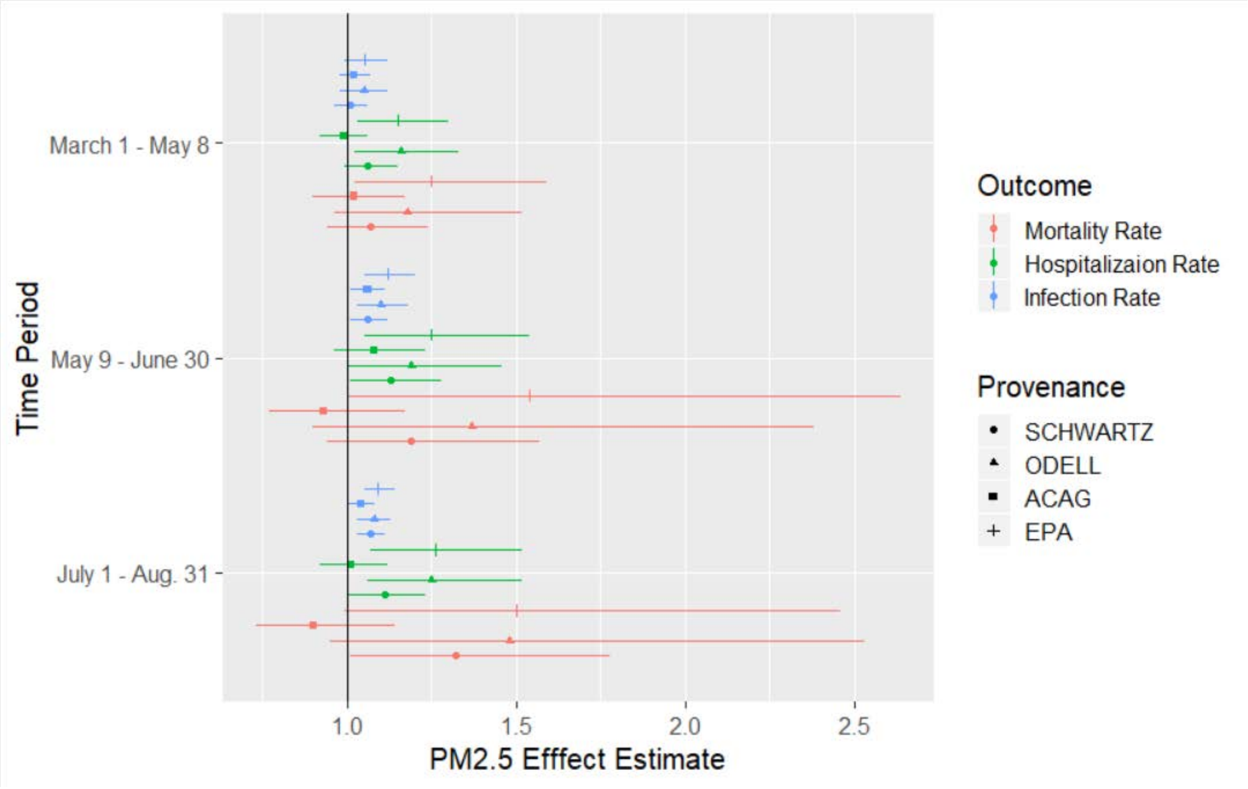


Figure S3: PM2.5 effect estimate during 3 distinct time periods
